# SARS-CoV-2 antibody seroprevalence in Tbilisi, the capital city of country of Georgia

**DOI:** 10.1101/2020.09.18.20195024

**Authors:** Tengiz Tsertsvadze, Lana Gatserelia, Marine Mirziashvili, Natia Dvali, Akaki Abutidze, Revaz Metchurtchlishvili, Carlos el Rio, Nikoloz Chkhartishvili

## Abstract

**Background:** Georgia timely implemented effective response measures, with testing, contact tracing and isolation being the main pillar of the national response, achieving the lowest cumulative incidence of SARS-CoV-2 in the European region.

**Methods:** We conducted a survey to estimate SARS-CoV-2 IgG antibody seroprevalence among adult residents of capital city of Tbilisi (adult population: 859,328). Participants were recruited through respondent driven sampling during May 18-27, 2020. Blood specimens were tested for SARS-CoV-2 IgG antibodies using commercially available lateral flow immunoassay (COVID-19 IgG/IgM Rapid Test Cassette, Zhejiang Orient Gene Biotech). Crude seroprevalence was weighted by population characteristics (age, sex, district of Tbilisi) and further adjusted for test accuracy.

**Results:** Among 1,068 adults recruited 963 (90.2%) were between 18 and 64 years-old, 682 (63.9%) women. 176 (16.5%) reported symptoms indicative of SARS-CoV-2 infection occurring in previous three months. Nine persons tested positive for IgG: crude seroprevalence: 0.84%, (95% CI: 0.33%-1.59%), weighted seroprevalence: 0.94% (95% CI: 0.37%-1.95%), weighted and adjusted for test accuracy: 1.02% (95% CI: 0.38%-2.18%). The seroprevalence estimates translate into 7,200 to 8,800 infections among adult residents of Tbilisi, which is at least 20 times higher than the number of confirmed cases.

**Conclusions:** Low seroprevalence confirms that Georgia successfully contained spread of SARS-CoV-2 during the first wave of pandemic. Findings also suggest that undocumented cases due to asymptomatic or very mild disease account for majority of infections. Given that asymptomatic persons can potentially spread the virus, test and isolate approach should be further expanded to control the epidemic.

## Introduction

Georgia, a small Eastern European country (population 3.7 million), rapidly responded to the threat of severe acute respiratory syndrome coronavirus 2 (SARS-CoV-2) infection. Early measures initiated in January-February 2020 included the approval of the emergency response plan on January 28; initiation of a communication campaign; stockpiling of personal protective equipment and establishing diagnostic capacity with real time reverse-transcriptase polymerase chain reaction (RT-PCR). Strict lockdown measures were implemented after the first case was diagnosed on February 26, including closure of schools and universities (March 2), national borders (March 18), restaurants and retail stores (except groceries and pharmacies, March 20), suspension of public transport (March 31). A state of emergency was declared on March 21 to enforce social distancing and isolation.^1^

Testing, contact tracing and isolation has been the main pillar of the national response, with all contacts of confirmed cases isolated for 14 days at one of the 84 hotels transformed to quarantine zones. Sixteen “fever” centers were established in the country for initial triage and RT-PCR diagnostics of suspected cases. Confirmed cases were transferred to one of the 12 dedicated hospitals. At the time of conducting survey (May 18-27) a cumulative 735 cases were reported and 12 died.^2^ As of September 12, 2020 a total of 2,075 cases were confirmed in the country with a cumulative incidence of 51.9 per 100,000 population, which the lowest rate in the European region.^3^

We conducted a survey to estimate SARS-CoV-2 antibody seroprevalence among adult residents of capital city of Tbilisi (adult population: 859,328).

## Methods

This was cross-sectional study of adult residents of Tbilisi. Eligibility criteria included age ≥ 18 years old, no confirmed SARS-CoV-2 infection and no current symptoms consistent with COVID-19.

Participants were recruited through respondent driven sampling during May 18-27, 2020. Sampling initiated with convenience sample of 25 individuals (seeds) with diverse sociodemographic characteristics representing all districts of Tbilisi. Additional 25 seeds were selected on day 5 to bolster recruitment. Each participant was allowed to recruit only 3 peers.

Consenting participants completed brief interview to elicit information about demographic characteristics, recent history of symptoms consistent with COVID-19, contact with confirmed or suspected case, employment in healthcare institution and history of international travel. Blood specimens were collected at Georgia’s referral institution for infectious diseases – Infectious Diseases, AIDS and Clinical Immunology Research Center. Those not able to come to the site were offered in-home blood collection. Specimens were tested for SARS-CoV-2 antibodies using commercially available lateral flow immunoassay (COVID-19 IgG/IgM Rapid Test Cassette, Zhejiang Orient Gene Biotech).

Crude seroprevalence was calculated as the proportion of survey participants testing positive for IgG antibodies. Crude estimate was weighted by population characteristics (age, sex and Tbilisi district) based on 2014 census data.^4^ Weighted estimate was further adjusted for test accuracy using published data from an independent validation study that showed sensitivity of 69% for IgM and 93.1% for IgG, specificity of 100% for IgM and 99.2% for IgG.^5^

Because of low sensitivity of IgM along with concerns related specificity and its rapid decay,^6,7^ our study focused on IgG as stronger marker of past infection.

Study was approved by the Institutional Review Board of the Infectious Diseases, AIDS and Clinical Immunology Research Center (OHRP #: IRB00006106). Informed consent was obtained from all participants.

## Results

A total of 1,068 adults were recruited excluding seeds. Ninety percent (n = 963) were between 18 and 64 years-old and 682 (63.9%) women (Table 1). Distribution of our sample significantly deviated from the population structure of Tbilisi including by age, sex and district justifying the need for adjustment on this parameters (Table 1).

**Table 1.** Distribution of study sample and adult population of Tbilisi by age, sex and district.

|  | Sample |  | Population of Tbilisi |  |
| --- | --- | --- | --- | --- |
|  | N | % | n | % |
| **Total** | 1068 | 100% | 859,328 | 100% |
| **Age categories** |  |  |  |  |
| 18-64 | 964 | 90.3% | 726,484 | 84.5% |
| 65+ | 104 | 9.7% | 132,844 | 15.5% |
| **Sex** |  |  |  |  |
| Women | 682 | 63.9% | 485,693 | 56.5% |
| Men | 386 | 36.1% | 373,635 | 43.5% |
| **Districts of Tbilisi** |  |  |  |  |
| Gldani | 115 | 10.8% | 135,212 | 15.7% |
| Didube | 70 | 6.6% | 55,205 | 6.4% |
| Vake | 182 | 17.0% | 88,076 | 10.2% |
| Isani | 62 | 5.8% | 96,787 | 11.3% |
| Krtsanisi | 23 | 2.2% | 29,960 | 3.5% |
| Mtatsminda | 42 | 3.9% | 39,054 | 4.5% |
| Nadzaladevi | 79 | 7.4% | 119,684 | 13.9% |
| Saburtalo | 339 | 31.7% | 108,392 | 12.6% |
| Samgori | 91 | 8.5% | 135,056 | 15.7% |
| Chughureti | 65 | 6.1% | 51,902 | 6.0% |

Sixty nine (6.5%) reported contact with either suspected or confirmed case and 186 (17.4%) persons have been working in healthcare institution other than clinics/hospitals treating COVID-19 patients.

Overall 176 (16.5%) participants reported symptoms indicative of SARS-CoV-2 infection occurring in previous three months, including 38 (3.6%) reporting fever > 38°C with any combination of other symptoms (cough, shortness of breath, fatigue, sore throat, rhinorrhea, loss of smell/taste), 108 (10.1%) had fever between 37-< 38°C with any combination of other symptoms (cough, shortness of breath, fatigue, sore throat, rhinorrhea, loss of smell/taste), and 30 reported any combination symptoms (cough, shortness of breath, fatigue, sore throat, rhinorrhea, loss of smell/taste) without fever. None of participants were hospitalized for these symptoms and no case of clinically/radiologically confirmed pneumonia was reported. Additional 164 (15.4%) participants reported symptoms occurring > 3 months ago.

Nine persons tested positive for IgG with crude seroprevalence of 0.84% (95% CI: 0.33%-1.59%) (estimated 7,200 adults). Weighted seroprevalence was 0.94% (95% CI: 0.37%-1.95%) (estimated 8,100 adults), which increased to 1.02% (95% CI: 00.38%−2.18%) (estimated 8,800 adults) after adjusting for test accuracy. Persons reporting symptoms in previous three months had higher seroprevalence (4.08% vs. 0.52%, p=0.01) (Table 2).

**Table 2.** Seroprevalence of SARS-CoV-2 IgG antibodies in Tbilisi, Georgia.

|  |  |  | **Seroprevalence, % (95% CI)** |  |  |  |  |  |
| --- | --- | --- | --- | --- | --- | --- | --- | --- |
|  | **No.** | **No.**<br>**positive**<br>**for IgG** | **Unweighted** | **p**<br>**value** | **Weighted for**<br>**population**<br>**characteristics** | **p**<br>**value** | **Wieghted and**<br>**adjusted for test**<br>**accuracy** | **p value** |
| **Total sample** | 1068 | 9 | 0.84 (0.33-1.59) |  | 0.94 (0.37-1.95) |  | 1.02 (0.38-2.18) |  |
| **Age categories** |  |  |  |  |  |  |  |  |
| 18-64 | 964 | 7 | 0.73 (0.29-1.49) | 0.22 | 0.71 (0.22-1.72) | 0.23 | 0.77 (0.24-1.84) | 0.23 |
| 65+ | 104 | 2 | 1.92 (0.23-6.77) |  | 2.21 (0.18-8.79) |  | 2.39 (0.25-8.84) |  |
| **Sex** |  |  |  |  |  |  |  |  |
| Women | 682 | 6 | 0.88 (0.32-1.90) | 1.00 | 1.12 (0.31-2.84) | 0.59 | 1.21 (0.46-2.96) | 0.58 |
| Men | 386 | 3 | 0.78 (0.16-2.25) |  | 0.71 (0.10-2.45) |  | 0.77 (0.10-2.68) |  |
| **Contact with suspected or confirmed case** |  |  |  |  |  |  |  |  |
| Yes | 69 | 0 | 0.0 (0.00-5.21) | 1.00 | 0.0 (0.00-6.06) | 1.00 | 0.0 (0.00-5.96) | 1.00 |
| No | 999 | 9 | 0.90 (0.41-1.70) |  | 1.00 (0.37-2.15) |  | 1.07 (0.39-2.36) |  |
| **History of symptoms** |  |  |  |  |  |  |  |  |
| Symptoms in previous 3 months | 176 | 4 | 2.27 (0.62-5.72) | 0.046 | 3.78 (0.76-10.79) | 0.009 | 4.08 (0.83-11.60) | 0.01 |
| No symptoms in previous 3 months | 892 | 5 | 0.56 (0.18-1.30) |  | 0.48 (0.13-1.21) |  | 0.52 (0.13-1.38) |  |
| **History of international travel** |  |  |  |  |  |  |  |  |
| Travel in previous 3 months | 83 | 2 | 2.41 (0.30-8.64) | 0.15 | 1.71 (0.11-7.36) | 0.48 | 1.85 (0.14-7.57) | 0.42 |
| No travel in previous 3 months | 985 | 7 | 0.71 (0.29-1.46) |  | 0.89 (0.29-2.08) |  | 0.96 (0.30-2.29) |  |
| **Employment type** |  |  |  |  |  |  |  |  |
| Healthcare | 186 | 2 | 1.08 (0.13-3.83) | 0.66 | 1.97 (0.18-7.63) | 0.27 | 2.13 (0.21-8.08) | 0.26 |
| Non-healthcare | 882 | 7 | 0.79 (0.32-2.63) |  | 0.75 (0.25-1.72) |  | 0.81 (0.26-1.91) |  |

Among four IgG positive persons with previous symptoms, three had had fever < 38°C and one had loss of smell/taste without any other symptoms, in addition one person also had recent history of travel to EU country. None of 164 persons reporting symptoms > 3 months ago tested positive for antibodies.

No statistically significant differences were found in seroprevalence by age, sex, history of contact with suspected/confirmed case, history of international travel and employment in healthcare institution (Table 2).

Similar seroprevalence was found in sensitivity analyses when comparing age and sex stratified rates (weighted and adjusted for test accuracy): 0.88% (95% CI: 0.13-2.95) among men vs. 0.69% (95% CI: 0.15-1.97) among women (p=0.77) in the age group of 18-64 years old; 0.0% (95% CI: 0.0-6.38) among men vs. 3.67% (95% CI: 0.30-14.31) among women (p=0.30) in the age group of 65+ years.

Only one person (80 years old lady) had isolated IgM positive test result, who was tested for active infection with nasopharyngeal swab RT-PCR immediately after positive IgM and one week thereafter. Both RT-PCR tests returned negative.

## Discussion

The seroprevalence of 0.84% to 1.02% shown in our survey suggests that between 7,200 and 8,800 persons have been infected with SARS-CoV-2 in Tbilisi, which is at least 20 times higher than the number of confirmed cases. Our findings are similar to those of other seroprevelence studies,^8–12^ corroborating previous finding that undocumented infections account for majority of cases.^13^

Georgia has established robust response strategy based on extensive testing, rapid detection, contact tracing and isolation. RT-PCR testing for SARS-CoV-2 is widely available for symptomatic cases and their contact as well as for populations at high risk (such as healthcare workers). Therefore undocumented cases in the country are most likely due to asymptomatic or very mild infections. National surveillance data indicates that 11% of confirmed cases did not have any symptoms at the time of diagnosis with 6% remaining asymptomatic throughout the course of disease.^14^ This is lower than recent estimates suggesting that 15 to 45% of all SASR-CoV-2 infections are asymptomatic.^15,16^ Consequently we believe that difference in estimated and reported numbers in Georgia is due to silent transmission from presymptomatic and asymptomatic cases, which has been shown to account for more than 50% of transmissions.^17–19^

As it would be expected people with history of symptoms had higher seroprevalence. But it should be noted that none of these persons had severe disease with sub-febrile temperatures and mild symptoms. Another indirect measure that supports our conclusion that asymptomatic or mild infections have been driving transmission in Georgia is no excess mortality observed in the country during the first 6 months of 2020. Moreover, the total number of death registered between January 1 and June 30, 2020 was 7% lower compared to 2019 and 1% less than in 2018.^14^

It is surprising that none of contacts of suspected or confirmed case had IgG antibodies. Survey used a simple questionnaire, to obtain very basic information and no details are available about the timing and type exposure, e.g. use of facemask, distance between persons etc. Also, seroconversion may have been missed if testing was done shortly after potential exposure or if they had asymptomatic/very mild disease.^20^ At the same time underreporting of being in contact with infected person because of stigma cannot be ruled out,^21^ resulting in underestimation of antibody prevalence in exposed persons.

Healthcare workers worldwide have been disproportionately affected by the pandemic.^22^ However, in our survey employees of healthcare institutions did not show higher seroprevalence that can be explained by the fact that none of them represented clinics/hospitals directly involved in care of COVID-19 patients.

There is no consistent data related to gender-specific susceptibility to infection. In our study especially older women, tended to have high seroprevalence, but the difference did not reach statistical significance. Recent report from china showed age-dependent increase in incidence among women, with greater difference observed among females aged 50-69 compared to males.^23^ On contrary, analysis of data from 10 European countries showed greater susceptibility among women of working age.^24^ Determining gender-based differences is beyond the scope of present work, but this warrants attention as it seems that social and economic aspects may play a significant role.

Although initial spread of the epidemic in Georgia was heavily associated with importation of cases from high incidence areas, recent history of international travel was not associated with higher seroprevalence in our study. The Government of Georgia closed national borders on March 18 within 3 weeks of first case was confirmed in the country, however given the high reproduction number SARS-CoV-2 of up to 4.^25^ this short period of time was sufficient to trigger local transmission.

Our study focused on IgG antibodies only. In addition to well-known drawbacks of determining IgM in terms of low sensitivity of test and rapid decay of antibodies, cross-reactivity with rheumatoid factor antibodies has been documented.^26^ In our survey only one person had isolated IgM antibodies, which we believe was a false positive result as active infection was not confirmed by repeated RT-PCR. In addition this was 80 years old person and age related increase in autoantibodies can be observed in healthy elderly.^27^

We acknowledge that the prevalence shown in our study might be biased because of limitations related to study design and test-accuracy. Survey was conducted during lockdown and applying probability-based random sampling was not feasible. Instead we used responded driven sampling, which is superior method over simple convenience sampling.^28^ In addition we offered in-home blood collection to facilitate enrollment in the study with almost 30% of participants opting for this options. However, bias could not be eliminated as we observed underrepresentation of certain population sub-groups, which was addressed by weighting our results by age, gender and district to match the population of Tbilisi. Another potential source of selection of bias is that persons with history of symptoms or contact with confirmed cases may have been more likely to participate to seek antibody confirmation of their potential exposure to SARS-CoV-2. We adjusted our estimates for test accuracy using data from independent validation study,^5^ however possibility of cross-reaction with other human coronaviruses remains a concern and cannot be ruled out.^^29–31^^

Despite these limitations study provides important data for informing public health action. First of all, our results highlight that Georgia’s response strategy succeeded in containing the epidemic. Seroprevalence of 1.02% is significantly lower than those reported from COVID-19 hotspot areas.^32^ Data also shows that despite strict confinement measures estimated number of infections was significantly greater than reported cases, likely resulting from silent transmission from asymptomatic or mild cases. Therefore, test and isolate approach along with social distancing, mask wearing, and general hygiene measures should be further expanded in the country to keep the epidemic under control. Repeated serosurveys, preferably using random sampling approach, are warranted to gain insights into pandemic trajectories.

## Data Availability

Data availability is restricted in accordance with national laws and regulations. Data are available from the Infectious Diseases, AIDS and Clinical Immunology Research Center for qualifying researchers. Data requests can be made at.

## Acknowledgement

Authors extend their gratitude to all participants for taking part in this survey.

